# Dissociation of white matter bundles in different recovery measures in post-stroke aphasia

**DOI:** 10.1101/2024.03.20.24304650

**Authors:** Alberto Osa García, Simona Maria Brambati, Amélie Brisebois, Bérengère Houzé, Christophe Bedetti, Alex Desautels, Karine Marcotte

## Abstract

**Background:** Post-stroke aphasia (PSA) recovery shows high variability across individuals and at different moments during recovery. Although diffusion biomarkers from the ventral and dorsal streams have demonstrated strong predictive power for language outcomes, it is still unclear how these biomarkers relate to the various stages of PSA recovery. In this study, we aim to compare diffusion metrics and language measures as predictors of language recovery in a longitudinal cohort of participants with PSA.

**Methods:** Twenty-four participants (mean age = 73 years, 8 women) presenting PSA were recruited in an acute stroke unit. Participants underwent diffusion MRI scanning and language assessment within 3 days (acute phase) after stroke, with a behavioral follow-up at subacute (10±3 days) and chronic phases (> 6 months). We used regression analyses on language performance (cross-sectional) and Δscores at subacute and chronic timepoints (difference between acute and subacute, and subacute and chronic respectively), with language baseline scores, diffusion metrics from language-related white matter tracts, lesion size and demographic predictors.

**Results:** Best prediction model of performance scores used axial diffusivity (AD) from the left arcuate fasciculus (AF) in both subacute (R^2^ = 0.785) and chronic timepoints (R^2^ = 0.626). Moreover, prediction of change scores depended on AD from left inferior frontal-occipital fasciculus (IFOF), in subacute stage (R^2^ = 0.5), and depended additionally on AD from right IFOF in the chronic stages (R^2^ = 0.68). Mediation analyses showed that lesion load of left AF mediated the relationship between AD from left AF and chronic language performance.

**Conclusion:** Language performance in subacute and chronic timepoints depends on the integrity of left AF, whereas Δscores of subacute and chronic phases depends on left IFOF, showing a dissociation of the white matter pathways regarding language outcomes. These results support the hypothesis of a functional differentiation of the dual-stream components in PSA recovery.

## INTRODUCTION

Aphasia is one of the most devastating post-stroke cognitive sequelae, with approximately 30% of stroke survivors presenting persistent language impairments (1). Despite its common prevalence, predicting outcomes for individuals with acute post-stroke aphasia (PSA) remains challenging due to heterogeneity of recovery patterns at the individual level (2). Recent reviews agree that lesion-related factors, such as initial aphasia severity, lesion size, and affected structures (3–5) are more relevant predictors of outcomes than demographic factors (e.g., age, education). Along with improvement of neuroimaging techniques and statistical approaches (6–9), modeling of PSA recovery has become an important goal in aphasia research. Two aspects of recovery have increasingly received attention: how to measure the recovery phenomenon and what brain regions and structures are associated with this recovery.

The time-bracketing of recovery is crucial when investigating longitudinal aphasia recovery, but its definition varies among researchers. The lack of precise definition for early and late recovery periods in literature may impact the interpretation of neuroimaging measurements, particularly in capturing temporal pathophysiological changes in early stages of recovery (10,11), or ongoing brain vascular damage in later stages (12). The groundbreaking neuroimaging findings by Saur and colleagues (12) suggest three important timepoints to define recovery, namely the acute (14–16), subacute (17,18), and chronic (18,19) phases. When analyzing the predictors of aphasia recovery, the choice of the outcome measures is also a complex question because most severity measures are weighted by initial severity. Cross-sectional language scores reflect performance at specific time points, while change scores capture performance dynamics between two timepoints, representing, namely, early (acute-subacute) or late (subacute-chronic) recovery phases (10–12).

Recent theories on aphasia recovery underscore the importance of quiescent areas in the brain’s post-stroke neuroplasticity (21). Significant recovery occurs both in cases of severe language impairment (22,23) and in lesions affecting core cortical language areas (3,13). This suggests there could be a post-stroke reorganization of the language network, which may be mediated by white matter bundles. Namely, the integrity of bundles from the dorsal stream, such as arcuate fasciculus (AF) and superior longitudinal fasciculus (SLF), has been associated with a better outcome in both early and chronic phases (15,16,24,25). Other studies have highlighted the significant role of ventral stream bundles, such as inferior fronto-occipital fasciculus (IFOF), inferior longitudinal fasciculus (ILF) and uncinate fasciculus (UF), in different phases of aphasia recovery (18,26). For example, Zavanone and colleagues observed a transition of the relation between language outcomes and lesioned voxels at white matter bundles, shifting from more anterior-frontal in early phases of recovery to posterior-temporal in later stages (18). This shift was interpreted as a progression from solely anterior parts of the dorsal stream to a convergence of dorsal and ventral streams in later recovery. It was hypothesized that the involvement of both dorsal and ventral streams is crucial for an optimal recovery. However, it remains unclear whether if overall recovery is related to preserved bundles from both streams, and whether this relation is consistent across different recovery phases (21,25).

This study has two objectives. Firstly, we aim to compare language outcomes between early and chronic phases in PSA and quantify the magnitude of change between these measures. Building on our previous research (24), we anticipate an improvement in language abilities over time, albeit this magnitude being similar between early and chronic phases. Secondly, we seek to explore the distinct roles of dorsal and ventral stream bundle integrity in predicting language performance and changes at various post-stroke timepoints. We hypothesize that if there is a specific phase-pathway association, we should observe a dissociation between pathway measures and language outcomes at different timepoints.

## METHODOLOGY

### 1. Participant recruitment and procedure

We recruited 39 participants with aphasia resulting from a first ischemic stroke in the left middle cerebral artery territory at an acute care hospital affiliated with the Centre de recherche du Centre intégré universitaire de santé et de services sociaux du Nord-de-l’Île-de-Montréal (CIUSSS NIM). Eligibility, confirmed by a neurologist, required proficient French and/or English skills before the stroke, with exclusion criteria including left-handedness, major psychiatric or developmental disorders, severe perceptual deficits, or other major neurological conditions. Following the protocol outlined by Saur and colleagues (13), language assessments and MRI scans were conducted within 3 days (acute), 7-15 days (early subacute), and at least 6 months (chronic) post-stroke. Fifteen participants completed only one or two assessments and/or withdrew from the study. Among the remaining participants, only 21 agreed to a subacute MRI scan, and of these, only 15 consented to a chronic MRI scan. To ensure consistency across timepoints, we utilized data from acute MRI scans alongside language data from all three timepoints, resulting in a final sample of 24 participants (mean age= 70 years, SD= 13; 8 women). Ethics approval was granted by the CIUSSS NIM ethics committee (#MP-32-2018-1478), and written informed consent was obtained from all participants or their legally authorized representatives.

### 2. Language assessment and variable creation

Based on previous studies (22,24), we calculated composite scores (CS), representing cross-sectional language performance, consisting of three sub-scores: comprehension, repetition and naming. The comprehension sub-score was composed of the word-sentence comprehension score from the Montreal-Toulouse aphasia battery (27) and the score of the revised short version of the Token Test (28). The repetition sub-score was composed of the word repetition and sentence repetition tasks from the MT-86 (27). The naming sub-score was composed of the semantic fluency score of the Protocole Montréal d’Évaluation de la Communication (29), and either the score of the Dénomination orale d’images (30) for participants speaking French or the score of the Boston Naming Test (31) for participants speaking English. All these tests are extensively used in both English and French speech pathology practice. The total score of each sub-score was reported on 10, resulting in CS maximum = 30. Three CS’s were calculated for each participant: acute (CS_1_), subacute (CS_2_), and chronic (CS_3_) scores.

Then we calculated the change of these scores in time. Change measures included early change (ΔCS_1_-_2_ = CS_2_ – CS_1_) and late change (ΔCS_2_-_3_ = CS_3_ – CS_2_) scores. Additionally, we computed relative recovery scores, with early relative change score (rΔCS_1_-_2_ = (CS_2_ – CS_1_)/ CS_1_) and late relative change score (rΔCS_2_-_3_ = (CS_3_ – CS_2_)/ CS_2_), emphasizing the gains of individuals with more severe impairments at baseline (32).

### 3. Neuroimaging procedure

#### 3.1. Image acquisition

An MRI scan including a diffusion sequence was performed on each participant on the day of the initial language assessment. The MRI images were acquired with a Skyra 3T scanner (Siemens Healthcare, USA). A high-resolution 3D T1-weighted image was acquired (TR = 2200 ms, TE = 2.96 ms, TI = 900 ms, FOV = 250 mm, voxel size = 1×1×1 mm3, matrix = 256156 cl:393256, 192 slices, flip-angle = 8°), in a Magnetization Prepared Rapid Gradient Echo (MP-RAGE) sequence. The MRI diffusion-weighted images had the following parameters: 65 images with non-collinear diffusion direction in b = 1000 s/mm2, posterior-anterior acquisition (TR = 11000 ms, TE = 86 ms, field of view = 230 mm, voxel resolution = 2 156 × 2 × 2 mm3, flip angle = 90°, bandwidth = 1700, EPI factor = 67), and two T2-weighted images with b = 0 s/mm2, one being a posterior-anterior acquisition, the other an anterior-posterior acquisition (time of acquisition = 12 min 30 s).

#### 3.2. DWI pre-processing and metric extraction

All pre-processing corrections and diffusion measure extractions were completed with the automated and reproductible pipeline Tractoflow (33). The procedure was supervised by three co-authors (CB, BH and SMB). First, we performed noise correction, using Marchenko-Pastur principal component analysis through MRtrix3 (34); subject movement and induced distortion correction through the FSL package (35); and a correction for N4 bias through ANTs package (36). Then, we performed the DTI metric extraction using DIPY (37), and T1-weighted image registration using non-linear SyN through ANTs (36). We used the Atlas Based Segmentation profile of TractoFlow version 2.3.0, using Freesurfer anatomical images in the within-subject template space, particularly recommended on pathological data (33). Once the tractogram was reconstructed, diffusion measures, namely fractional anisotropy (FA), mean diffusivity (MD) and axial diffusivity (AD), were extracted from bundles of ventral stream (IFOF, ILF and UF) and the dorsal stream (AF and SLF) bilaterally.

Finally, we performed a semi-automated segmentation of each brain lesion in acute phase using *Clusterize* (38). This toolbox (http://www.medizin.uni-tuebingen.de/kinder/en/research/neuroimaging/software/), running with SPM12 under Matlab (The Mathworks, Inc., Natick, MA), has previously shown to have a good reliability in lesion demarcation in acute post-stroke patients (De Haan et al., 2015). Clusters of hypointense voxels were first identified on MD maps (set with default parameters). Then, cluster(s)-of-interest were manually selected and adjusted to fit the lesion in each slice and adjusted (if needed) with MI-brain software (Imeka Solutions Inc.), with the help of MD maps and b0 DWI maps by a study team member (BH). Finally, each lesion file was counter-verified by another team member (SMB) with the help of MD maps and b0 DWI maps. Both raters are experiences in lesion delineation and were blinded to the participant’s identity. Volume of the lesions and intracranial volumes were extracted in mL and corrected for intracranial volume. Lesion loads for each left-hemisphere tract were also calculated using the mask of the lesion and the parceled area of the tracts, with a correction for tract volume afterwards.

## 3.3. Statistical proceeding

To evaluate the impact of time on language performance scores, we conducted a one-way ANOVA on all CS, and paired tests on the difference of ΔCS and rΔCS in both early and late recovery phases. Additionally, we examined whether receiving rt-PA affected any language outcome. Next, we identified diffusion predictors for inclusion in regression models. In the subacute phase, the dependent variables were CS_2_, ΔCS_1-2_ and rΔCS_1-2_. In the chronic phase, the dependent variables were CS_3_, ΔCS_2-3_ and rΔCS_2-3_. All analyses were performed using SPSS (version 28.0.1.0). Model selection is illustrated in Figure 1 and involved the following steps:

1) For each stream (dorsal and ventral stream), we conducted backwards regressions, entering the diffusion measures of each representative tract (dorsal stream: AF, SLF; ventral stream: ILF, IFOF, UF) from both hemispheres. Collinearity was monitored, and predictors were retained based on Variance Inflation Factor (VIF) and Condition Indices (CI). Predictors were excluded if they exhibited a correlation > 0.6 with another predictor in the same final model, showing a VIF > 3 or CI > 30. In the case of collinearity, the predictor with the higher standardized beta coefficient was retained.
2) We conducted nested linear regressions using retained predictors from step 1, incorporating lesion, language, and demographic measures. Each regression comprised a null model with only diffusion measures predicting each dependent variable, and a complete model with diffusion measures and covariates (age, education, lesion size, and the CS of the timepoint immediately prior to the studied timepoint). Subacute models included CS_1_, while chronic models had two versions, one with CS_1_ and another with CS_2_ as the baseline score. This comparison aimed to evaluate the relationship between acute (initial), subacute, and chronic scores, considering an expected larger change in the early weeks of recovery.
3) A-posteriori mediation analyses were conducted to assess the influence of lesion loads in the left hemisphere language bundles for each successful measure in the nested models. Each mediation analysis involved a diffusion measure as the predictor and language measures as responses.

**Figure 1.**
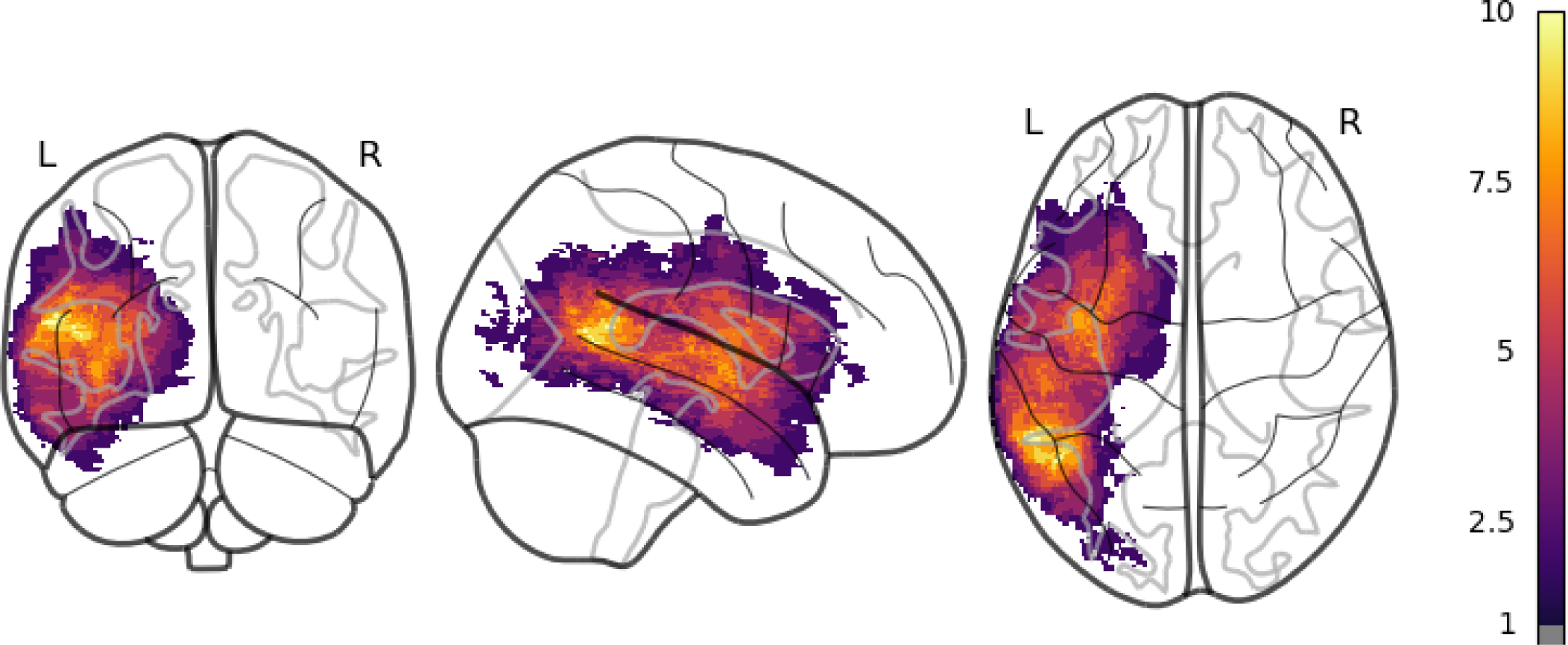
Lesion overlay map of participants in the study. Color scale indicates minimum number of participants with the same location for lesion voxels.

**Figure 2.**
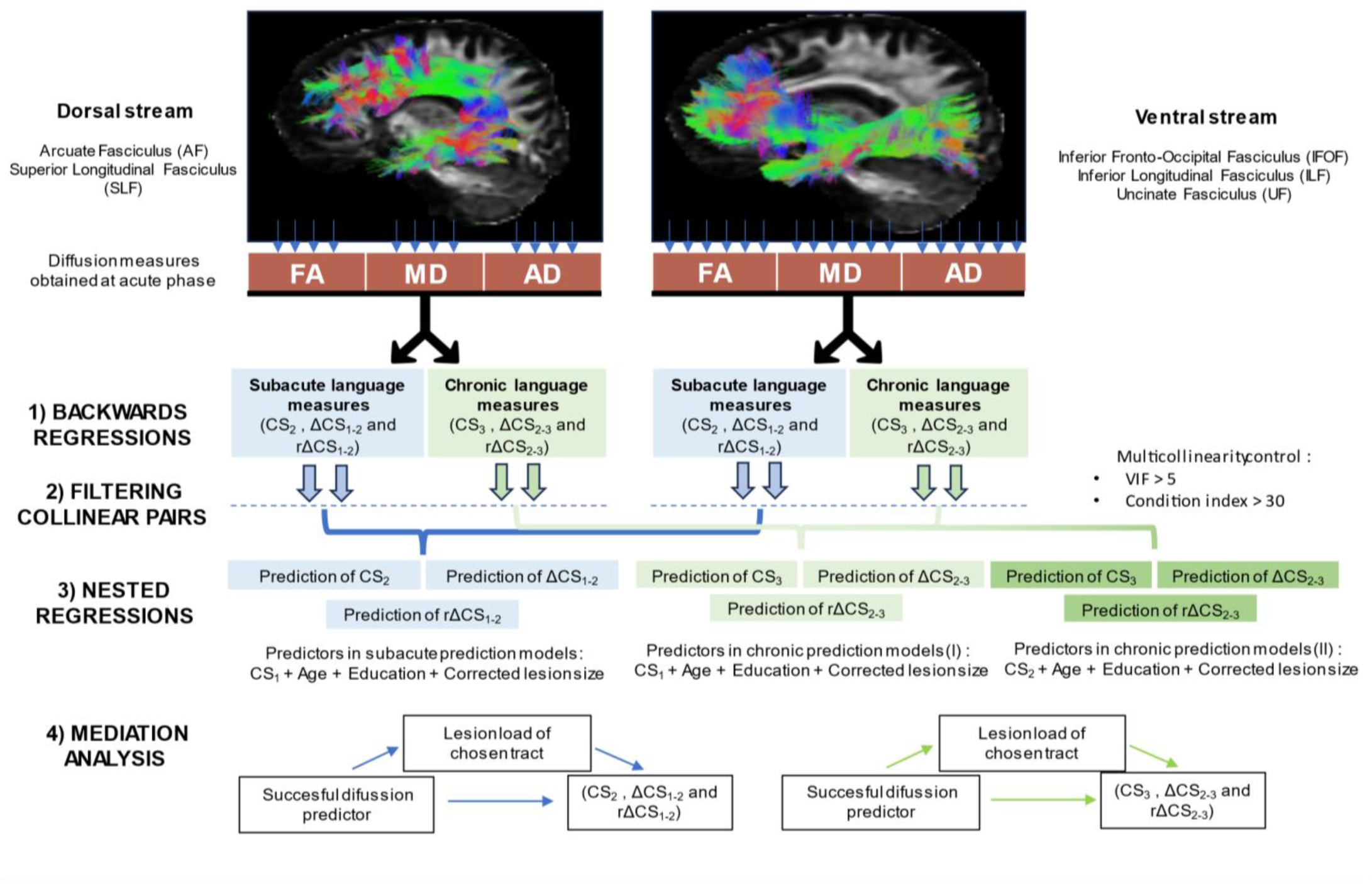
Procedure used for variable selection and model construction. CS= Composite score; FA= Fractional Anisotropy; MD= Mean Diffusivity; AD: Axial Diffusivity.

**Figure 3.**
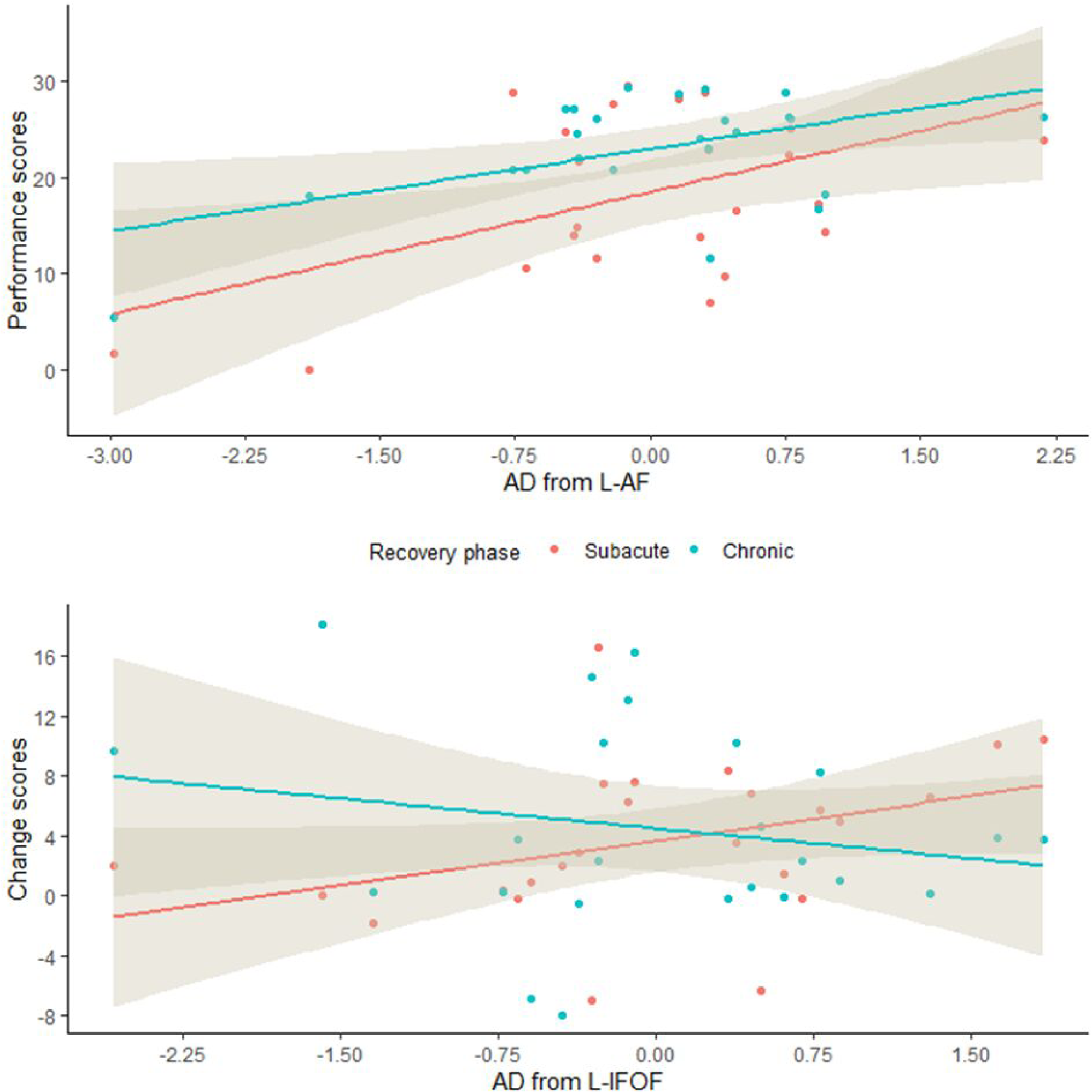
Scatterplots with the relation of predictive diffusion (AD from L-AF and AD from L-IFOF) measures and the language outcomes for which they had significant coefficients. X-axis variables are shown scaled.

## RESULTS

Demographic, clinical characteristics and CS are reported in Table 1. Friedman’s ANOVA analysis across the three CS was significant (χ^2^ = 28.2, p<0.001, effect size Kendall’s W= 0.587). Conover post-hoc comparisons, with Holm correction applied, showed a difference between the acute and subacute timepoints (T [46] =2.32, p=0.025) and between the subacute and chronic timepoints (T [46] = 5.297, p<0.001). No difference in change measures were found between ΔCS_1-2_ and ΔCS_2-3_ (t [23] = −0.724, p=0.477) or between rΔCS_1-2_ and rΔCS_2-3_ (W=169, p=0.6). Participants that received rt-PA had a higher CS_1_ (t [22] = −2.225, p=0.037, Cohen’s d=0.45), CS_2_ (W = 37, p=0.047, Cohen’s d=0.23) and CS_3_ (W = 36, p=0.041, Cohen’s d=0.23) than their counterparts, but there was no difference for ΔCS_1_-_2_ (W= 72, p = 0.9), and ΔCS_2_-_3_ (W =96, p=0.16). Visualization of the trajectory of sub-scores, CS and change scores can be found in the supplementary material.

**Table 1.** Demographic and clinical characteristics of participants.

| Variable | Mean ± SD | Range |
| --- | --- | --- |
| Sex (M/F) | 16 M; 8 F |  |
| Age (years) | 70 ± 13 | (47, 95) |
| Education (years) | 11.3 ± 5.5 | (6, 26) |
| Lesion site (F/T/P/SC)* | 11/10/10/12 |  |
| Lesion size (mL) | 24.75 ± 24.7 | (0.16, 84.42) |
| Thrombolysis received (Y/N) | 13/11 |  |
| Time Stroke - Assessment<sub>1</sub> (days) | 2 ± 0.8 | (1, 3) |
| Time Stroke - Assessment<sub>2</sub> (days) | 10 ± 2.1 | (7, 15) |
| Time Stroke - Assessment<sub>3</sub> (days) | 270 ± 60 | (180, 500) |
| Speech / Language therapy (Y/N) <sup>†</sup> | 21/3 |  |
| CS<sub>1</sub> | 15.3 ± 9.13 | (0, 28.5) |
| CS<sub>2</sub> | 20.3 ± 8.5 | (0, 29.5) |
| CS<sub>3</sub> | 25.8 ± 5.3 | (5.3, 29.7) |
| ΔCS<sub>1-2</sub> | 3.8 ± 5.2 | (-9.9, 16.5) |
| ΔCS<sub>2-3</sub> | 5.07 ± 5.8 | (-0.56, 18.12) |
| rΔCS<sub>1-2</sub> <sup>‡</sup> | 22.37 / 3.8 + 101.1 / 3.11 | (- 46.1, 372.7) / (- 4.52, 6.61) |
| rΔCS<sub>2-3</sub> <sup>‡</sup> | 13.55 / 3.26 + 59.7 / 2.46 | (-2.84, 219.36) / (-1.76, 6.08) |

\*F= Frontal; T= Temporal; P= Parietal; SC= Subcortical
<sup>†</sup> Y = Received speech/language therapy; N = Did not receive therapy. Minimum reported = one month with three weekly sessions with a speech-language therapist, maximum reported duration = 12 months.
<sup>‡</sup> Raw relative change scores / Relative change scores after Box-Cox transformation to address high heteroscedasticity. Analyses were performed using transformed values.

## DIFFUSION PREDICTORS

Coefficients of successful diffusion predictors after backwards regressions are reported in Table 2. Regarding the prediction of performance measures (i.e., the CS), only measurements of the left AF from the dorsal stream were significant in both the subacute and chronic recovery phases (L-AF_AD_ and L-AF_FA_, for both CS_2_ and CS_3_). From the ventral stream, only L-UF_MD_ was significant predicting CS2. As for the prediction of absolute change scores, only measures from the ventral stream survived the regressions. Namely, L-IFOF_AD_ could well predict ΔCS_1-2,_ whereas L-UF_MD_, L-IFOF_AD_ and R-IFOF_AD_ could well predict ΔCS_2-3_. Regarding the prediction of relative change scores, L-IFOF_AD_ was the only successful predictor.

**Table 2.** Results from backwards regression models for each dependent variable. Only predictors with significant coefficients at α=0.05 and individual R^2^ obtained from part correlations are reported. Highly collinear predictor pairs were excluded.

|  | Subacute recovery phase |  |  |  |  |  |  |  |  | Chronic recovery phase |  |  |  |  |  |
| --- | --- | --- | --- | --- | --- | --- | --- | --- | --- | --- | --- | --- | --- | --- | --- |
| | CS<sub>2</sub> | | | $\Delta CS_{1,2}$ | | | $r\Delta CS_{1,2}$ | | | CS<sub>3</sub> | | | $\Delta CS_{2,3}$ | | |
| | Predictor | R<sup>2</sup> | $\beta_s$ | Predictor | R<sup>2</sup> | $\beta_s$ | Predictor | R<sup>2</sup> | $\beta_s$ | Predictor | R<sup>2</sup> | $\beta_s$ | Predictor | R<sup>2</sup> | $\beta_s$ |
| Dorsal stream | L-AF<sub>AD</sub> | 0.275 | 0.524 |  |  |  |  |  |  | L-AF<sub>AD</sub> | 0.266 | 0.516 |  |  |  |
|  | L-AF<sub>FA</sub> | 0.198 | 0.445 |  |  |  |  |  |  | L-AF<sub>FA</sub> | 0.192 | 0.438 |  |  |  |
| Ventral stream | L-UF<sub>MD</sub> | 0.169 | 0.411 | L-IFOF<sub>AD</sub> | 0.27 | 0.522 | L-IFOF<sub>AD</sub> | 0.168 | 0.41 |  |  |  | L-UF<sub>MD</sub> | 0.186 | -0.431 |
|  |  |  |  |  |  |  |  |  |  |  |  |  | L-IFOF<sub>AD</sub> | 0.33 | -0.81 |
|  |  |  |  |  |  |  |  |  |  |  |  |  | R-IFOF<sub>AD</sub> | 0.193 | 0.5 |

## COMPLETE REGRESSION MODELS

R^2^ values and standardized coefficients in all models can be found in Table 3. In summary, complete models improved significantly the variance explained by only-diffusion-predictors models in the prediction of CS_2_, CS_3_ using CS_2_, and all change scores. First, whenever CS was used as predictor it resulted being significant in the final model, except for the prediction models of ΔCS_2-3,_ whichever CS was used as baseline (with CS_1_, β = −0.495, p=0.077; with CS_2_, β = −0.488, p=0.104). Among diffusion predictors from the dorsal stream, we have only tested models with L-AF_AD,_ excluding L-AF_FA_, since two predictors from the same bundle and hemisphere could implicate high collinearity. L-AF_AD_ resulted significant in the prediction of CS_3_ using CS_1_ as baseline (β = 0.502, p=0.03). Among diffusion predictors from the ventral stream, L-IFOF_AD_ was the only predictor that proved to be significant in all tested models. L-IFOF_AD_ was significant in the prediction of ΔCS_1-2_ (β = 0.407, p=0.048), and ΔCS_2-3_ using CS_1_ (β = −0.65, p=0.026). No other covariates proved to be significant in the complete models. Only one model with a diffusion predictor (L-IFOF_AD_) was compared to a complete model in predicting relative change measure. The model showed a relatively small R^2^ (0.37), and no predictor was significant except for CS_1_. Chronic relative change score regressions had higher and similar R^2^ (R^2^ = 0.69 using CS_1_; R^2^ = 0.69 using CS_2_).

**Table 3.** Coefficients in prediction models of performance scores, raw change scores and relative change scores. Variables and elements in bold represent significant additions in the complete regression models at α=0.05.

| Model | CS<sub>2</sub> model |  | CS<sub>3</sub> model with CS<sub>1</sub> |  | CS<sub>3</sub> model with CS<sub>2</sub> |  |
| --- | --- | --- | --- | --- | --- | --- |
| Nested model | R<sup>2</sup> = 0.31 |  | R<sup>2</sup> = 0.26 |  | R<sup>2</sup> = 0.26 |  |
| Complete model | R<sup>2</sup> = 0.78 | **R<sup>2</sup> change**<br>**p < 0.001** | R<sup>2</sup> = 0.5 | R<sup>2</sup> change<br>p = 0.06 | R<sup>2</sup> = 0.62 | **R<sup>2</sup> change**<br>**p = 0.01** |
| Predictors | Stand.β | P value | Stand.β | P value | Stand.β | P value |
| Intercept |  | 0.85 |  | 0.425 |  | 0.36 |
| CS<sub>1</sub>/CS<sub>2</sub> | **0.53** | **0.015\*** | **0.465** | **0.019\*** | **0.81** | **0.004\*** |
| Age | - 0.11 | 0.44 | - 0.24 | 0.21 | - 0.19 | 0.29 |
| Education | - 0.20 | 0.15 | 0.1 | 0.61 | 0.275 | 0.15 |
| Lesion size | - 0.18 | 0.39 | 0.12 | 0.70 | 0.17 | 0.46 |
| L-AF<sub>AD</sub> | 0.11 | 0.54 | **0.50** | **0.034\*** | 0.41 | 0.077 |
| L-UF<sub>MD</sub> | 0.14 | 0.33 | - | - | - | - |
| Model | ΔCS<sub>1-2</sub> model |  | ΔCS<sub>2-3</sub> model with CS<sub>1</sub> |  | ΔCS<sub>2-3</sub> model using CS<sub>2</sub> |  |
| Nested model | R<sup>2</sup> = 0.27 |  | R<sup>2</sup> = 0.39 |  | R<sup>2</sup> = 0.39 |  |
| Complete model | R<sup>2</sup> = 0.50 | R<sup>2</sup> change<br>p = 0.123 | R<sup>2</sup> = 0.69 | **R<sup>2</sup> Change**<br>**p = 0.023\*** | R<sup>2</sup> = 0.68 | **R<sup>2</sup> Change**<br>**p = 0.029\*** |
| Predictors | Stand.β | P value | Stand.β | P value | Stand.β | P value |
| Intercept |  | 0.150 |  | 0.425 |  | 0.150 |
| CS<sub>1</sub>/CS<sub>2</sub> | **- 0.737** | **0.017\*** | -0.495 | 0.077 | **- 0.737** | **0.017\*** |
| Age | - 0.059 | 0.733 | 0.048 | 0.765 | - 0.059 | 0.733 |
| Education | - 0.224 | 0.290 | 0.08 | 0.660 | - 0.224 | 0.290 |
| Lesion size | - 0.341 | 0.222 | 0.102 | 0.663 | - 0.341 | 0.222 |
| L-IFOF<sub>AD</sub> | **0.407** | **0.048\*** | **- 0.65** | **0.026\*** | **0.407** | **0.048\*** |
| R-IFOF<sub>AD</sub> | - | - | 0.374 | 0.095 | - | - |
| L-UF<sub>MD</sub> | - | - | -0.01 | 0.965 | - | - |
| Model | rΔCS<sub>1-2</sub> model |  | rΔCS<sub>2-3</sub> model with CS<sub>1</sub> |  | rΔCS<sub>2-3</sub> model with CS<sub>2</sub> |  |
| Nested model | R<sup>2</sup> = 0.17 |  |  |  |  |  |
| Complete model | R<sup>2</sup> = 0.37 | R<sup>2</sup> Change<br>p = 0.26 | R<sup>2</sup> = 0.69 |  | R<sup>2</sup> = 0.62 |  |
| Predictors | Stand.β | P value | Stand.β | P value | Stand.β | P value |
| Intercept | - | 0.306 | - | 0.527 | - | 0.187 |
| CS<sub>1</sub>/CS<sub>2</sub> | - **0.705** | **0.039\*** | **-0.582** | **0.003** | -0.706 | 0.008 |
| Age | 0.092 | 0.638 | 0.157 | 0.318 | 0.142 | 0.336 |
| Education | - 0.221 | 0.351 | 0.236 | 0.161 | 0.075 | 0.663 |
| Lesion size | - 0.349 | 0.264 | 0.311 | 0.728 | 0.065 | 0.763 |
| L-IFOF<sub>AD</sub> | 0.276 | 0.219 | - | - | - | - |

## *A POSTERIORI*-MEDIATION ANALYSES

We only conducted mediation analyses between the lesion load of L-AF_AD,_ L-IFOF_AD_ and the response measures that were predicted in each model, since only those two measures proved to be significant in the full models. No mediation effect was found in the prediction of raw or relative change measures using L-IFOF_AD_. No mediation effect was either found in the prediction of CS_2_ using L-AF_AD_, however we found that the lesion load of AF worked as a mediator in the prediction of CS_3_ using L-AF_AD_ (Indirect effect = 21566, average mediation effect = 60%, CI= [4738.08, 44294.83], proportional CI= [0.17,1.4]; p = 0.008). In presence of the lesion load of AF, L-AF_AD_ was not a significant predictor of CS_3_ (β = 0.202, p=0.255), whereas the lesion load of AF proved to be significant (β = - 0.616, p=0.002). Visualization of the mediation effect of Lesion load on L-AF_AD_ regression on CS_3_ can be found in the supplementary material.

## DISCUSSION

In this study, we first compared the language outcomes from early and late phases of recovery and confirmed continuous improvement in language abilities. Consistent with seminal work in this field (23), we observed a similar degree of recovery in the early and late phases. Then, we explored the relationship between diffusion measures from the dual-stream system tracts and language outcomes in PSA. Results showed that acute AD from left AF predicted performance scores in subacute and chronic timepoints, while acute AD from left IFOF predicted early and late change scores, showing a negative trend in the chronic phase. Additionally, AD from right IFOF correlated positively with chronic change scores. Baseline language scores enhanced complete prediction models in all cases. All language outcomes (performance and change scores) could be well predicted by corresponding baseline language abilities, diffusion measures and covariates, as expected because of the relation between the outcome measure and the aphasia severity (i.e., CS) from the previous phases. Finally, results showed that lesion load only influenced the relationship between left AF and chronic cross-sectional outcomes. In summary, it was the nature of language measures (performance vs change scores) that differentiated the relationship between white matter tracts and language measures, rather than chronology of the language measures. The performance measure represents time-locked performance abilities whereas the change score represents the dynamic of performance between two timepoints. Thus, the present results could suggest that the associated tracts (i.e. left AF, left IFOF, left UF and right IFOF) may have different roles in the recovery trajectory. To our knowledge, this is the first time that such dissociation is reported.

L-AF_AD_ effectively predicted subacute and chronic performance scores in our PSA cohort, consistent with previous findings linking AF to optimal language abilities in PSA individuals, including naming (39), repetition (40,41), comprehension (39) and spontaneous speech (42,43). Other studies that have tried to relate change scores and AF measures have not obtained significant results either (44–46). Bae and colleagues (45) demonstrated a link between left AF diffusion changes (increased FA over time) and language score variations after 6 months, suggesting a positive impact of improved diffusion stability on language, albeit with different measurement timing than our study. However, change of diffusion measures requires a chronic measure, and we intended to investigate the utility of acute measures as predictors of chronic outcomes. However, this prediction was conditional on the overall integrity of the left AF, since lesion load seemed to be a mediator in that relation, echoing the importance of the left hemisphere dorsal pathway over its right counterpart in recovery (39,47). The lack of a similar effect on left IFOF measures and the exclusive association of left, not right, AF measures with language improvement emphasizes the sensitivity of this bundle to anatomical disruptions, consistent with other cohort studies (48).

Regarding ventral stream tracts, an exclusive relation to change scores was found in both recovery phases. Two results notably stand out: AD from right IFOF significantly predicted late changes, while AD from left IFOF changed the direction of its relation from positive (in early change prediction) to negative (in late change prediction). These two results may reflect the bilateral nature of the ventral pathway, particularly evident in the chronic phase, which would point to a requirement for bilateral ventral structures in long-term recovery. It should be noted, though, that this finding does not entirely support the need for right dorsal-stream structures, but rather reminds the inherent bilateral nature of the language abilities supported by the ventral streams. A recent study, exploring right hemisphere connectivity, precisely suggested a crucial role of these structures in later stages compared to early stages (17). Our results underscore this claim, highlighting the significance of language-related tracts from the dual-stream model. We consider important to remind how early phases (i.e., subacute performance, or early changes between subacute and acute) are indeed predictable, and our data suggest that higher stability of white matter bundles from both streams support language from the very beginning of the recovery process, being proof that early recovery can be well captured with neurobiological measures.

Our findings also primarily indicate a relationship between AD measures and language recovery, with FA only emerging as a predictor in one instance. Interestingly, almost all our regression analyses consistently identified AD as a better predictor. While AD has received less attention than FA in aphasia literature (15,39,46), recent work focusing on the late subacute phase in stroke survivors demonstrated its association with positive language outcomes (49). Higher AD in the language-related tracts could reflect higher stability, which also tends to persist over time. However, it is usual that AD rises in the acute phase of recovery, whereas FA remains stable for longer periods. As suggested by Moulton and colleagues (49), AD may provide valuable information on in the acute prognostic, since lower AD could indicate poor hyperacute recovery after stroke, though it may not decisively predict recovery in later stages.

Our study has certainly its limitations. Beyond the modest size of the sample, we relied on structural data to explore neural correlates of recovery, whereas connectivity measures can provide a deeper insight on the status of each tract and in each phase. Nevertheless, we consider that this study is valuable for the importance of reporting more than two timepoints in the same cohort including early acute measures, which has yielded important evidence about recovery in recent literature (2,13,44). Another limitation is the absence of specific cortical measures, although literature has vastly shown utility of both white and grey matter measures in predicting recovery (15,41). The choice of variables is also an important issue in aphasia recovery studies. While score changes are increasingly utilized in aphasia studies to represent language gains (2,17,21,44) they may not fully consider individual baseline abilities, potentially affecting the interpretation of results (32). Bae and colleagues (50) also addressed this concern but did not find significant results using proportional recovery measures. There may not be a linear relationship between relative recovery measures and diffusion measures (possibly due to heteroscedasticity), but we consider that behavioral studies should rightfully assess individual recovery patterns to enable meaningful group-level comparisons.

In conclusion, our data suggest that bundles from the dual-stream model may have a role in the recovery process, possibly either mediated by the language processes they support, or by a compensation that they confer for the remaining language abilities after the disruption of the language network. These hypotheses should be tested in future venues to further elucidate the nature of PSA recovery.

## Data Availability

Data is available upon contact and consideration with the corresponding author.

## Acknowledgments

We thank all participants, their families and health professionals from the clinical setting for their collaboration all along the project.

## Sources of Funding

This project was funded by a grant-in-aid from the Heart and Stroke Foundation (G-16-00014039 and G-19-0026212) as well as a project grant from the Canadian Institutes of Health Research (CIHR 470371) to K.M., A.D. and S.M.B. K.M. holds a Career Award from the “Fonds de Recherche du Québec – Santé”. A.O.G. holds a “Fonds de Recherche du Québec – Santé” scholarship.

## Disclosures

AD received research grants from Canopy Growth and Eisai, served on scientific advisory boards for Eisai, Jazz Pharma and Eisai, as well as honoraria from speaking engagements from Jazz Pharma and Paladin Labs. The remaining authors do not have any conflict of interest to declare.

## Notes

### Author Declarations

Ethical approval for this study was obtained by the Ethics Committee at CIUSSS-NIM ((#MP-32-2018-1478)

